# Process Mining/Deep Learning Model to Predict Mortality in Coronary Artery Disease Patients

**DOI:** 10.1101/2024.06.26.24309553

**Authors:** Negin Ashrafi, Armin Abdollahi, Greg Placencia, Maryam Pishgar

## Abstract

Patients with Coronary Artery Disease (CAD) are at high risk of death. CAD is the third leading cause of mortality worldwide. However, there is a lack of research concerning CAD patient mortality prediction; thus, more accurate prediction modeling is needed to predict the mortality of patients diagnosed with CAD. This paper demonstrates performance improvements in predicting the mortality of CAD patients. The proposed framework is a modification of the work used for the prediction of 30-day readmission for ICU patients with heart failure. Our framework demonstrates better performance with an Area Under the ROC Curve (AUC) score of 0.871 for the Neural Network (NN) model compared to traditional baseline machine learning models that we developed. Our framework uses the medical history of patients, the time related to the variables, and patients’ demographic information for prediction. This framework has the potential to be used by medical teams to make more accurate decisions for treatment and care for patients with CAD, increasing their life expectancy.

## 1. INTRODUCTION

Plaque buildup causes Coronary Artery Disease (CAD)[1]. Plaque, composed of cholesterol deposits, gradually accumulates inside the arteries, narrowing the walls of the coronary arteries that supply blood to the heart (called coronary arteries) [1]. Patients with CAD are at high risk of death. CAD causes approximately 610,000 deaths yearly (1 in 4 deaths) in the United States alone [2]. It is the third leading cause of mortality worldwide, associated with 17.8 million deaths annually [2]. The estimated annual cost of healthcare services for CAD in the United States exceeds 200 billion dollars [2]. Despite being a significant cause of death and disability, there are few accurate prediction models for the mortality of CAD patients.

Early prediction of CAD could help clinicians make better decisions about effectively allocating medical resources to save patients’ lives. Given the scarcity of existing literature predicting this critical outcome, current research must focus on developing more accurate models to predict the mortality of CAD patients and enhance their life expectancy.

A variety of models predict mortality for patients with critical diseases such as Paralytic Ileus [3] and diabetes [4]. Several models also identify risk factors related to the mortality of CAD patients [2]. However, there is a lack of prediction modeling for the mortality of CAD patients.

Besides machine learning, an innovative process mining/ deep learning framework has shown promise in predicting different health outcomes for ICU patients with a variety of diseases [4, 5]. Demographic information of patients, severity scores on admission day, and time information related to variables, called timed state samples, were produced through a process mining algorithm (Decay Replay Mining (DREAM)) [5], then fed into a Neural Network (NN) to predict healthcare outcomes. Significant improvements in AUC values were found compared to existing literature.

This work focuses on determining the mortality of patients with CAD using a process mining/ deep learning approach. Moreover, we compared results from the proposed approach with those of several machine learning models we developed. The proposed prediction framework demonstrates improved performance in predicting whether CAD patients die or not.

Section 2 of this paper summarizes the preliminaries needed throughout the paper. Section 3 introduces the methodology. Section 4 evaluates the approach by discussing the dataset, and process mining/ deep learning model architecture, and compares results against baseline models for predicting the mortality of CAD patients. Section 5 concludes by discussing future work.

## 2. Preliminaries

Notations in this section were adopted from [6].

### 2.1. Process Mining

Process mining analyzes processes using event logs. These methods are classified into process discovery, conformance checking, and process enhancement. In this work, a process discovery algorithm produced a process model commonly called Petri Net. Details on Process mining can be found in [7].

### 2.2. Petri Net

Petri Nets (PN) are a mathematical model for processes. PNs consist of a set of places graphically represented by circles and transitions (events) represented by rectangles. Process events are modeled by firing transitions corresponding to that event. Places can be marked (showing the current process state) or unmarked. Tokens (dots) in a circle represent place markings corresponding to that event. Transitions fire when all its input places (an input arrow to the transition) have the required number of tokens. When transitions fire, a predetermined number of tokens are removed from all its input places. Each output place (with an input arrow from the transition) also receives one or more tokens. Details about PNs are given in [7, 8, 9].

### 2.3. Event Log

Event *a ϵ A* instantaneously changes process state [4]. Event *a* can occur multiple times within an identified process, where *A* indicates the finite set of all identified events. *E* is an event instance, denoted by a vector with at least two attributes: the associated events’ name *a* and a corresponding timestamp, *τ. E* may contain resources. Two events cannot have the same timestamps [4]. A trace *g ϵ G* is a finite, ordered sequence of event instances [4].

### 2.4. Decay Replay Mining

Decay Replay Mining (DREAM) [5], is a process mining-based approach that can predict the subsequent process events. DREAM extends places discovered by the PN process model with time decay functions. During the replay of the event log on a PN, time decay functions use timestamp information as a parameter to generate timed state samples.

### 2.5. Timed State Samples

Timed state samples are generated after replaying an event log on the PN. The timed state sample contains time information that represents the state of the process [5].

## 3. Methodology

We explain the proposed method to predict the mortality of patients with CAD in this section. First, we describe feature selection. Next, we describe how Electronic Health Records (EHR) are converted to event logs. We then introduce methods to predict the mortality of patients with CAD using the DREAM algorithm. Finally, we compare the proposed process mining model results to those of baseline models. Pishgar et al proposed a similar framework in [3, 6, 10] to predict in-hospital mortality of ICU patients with Paralytic Ileus and to predict unplanned 30-day readmission of ICU patients with Heart Failure. Our framework modifies these frameworks to predict the mortality of CAD patients. However, the attributes and the architecture of the NN differ from those in [3, 6, 10]. The proposed prediction framework is outlined in Fig.1

**Figure 1:**
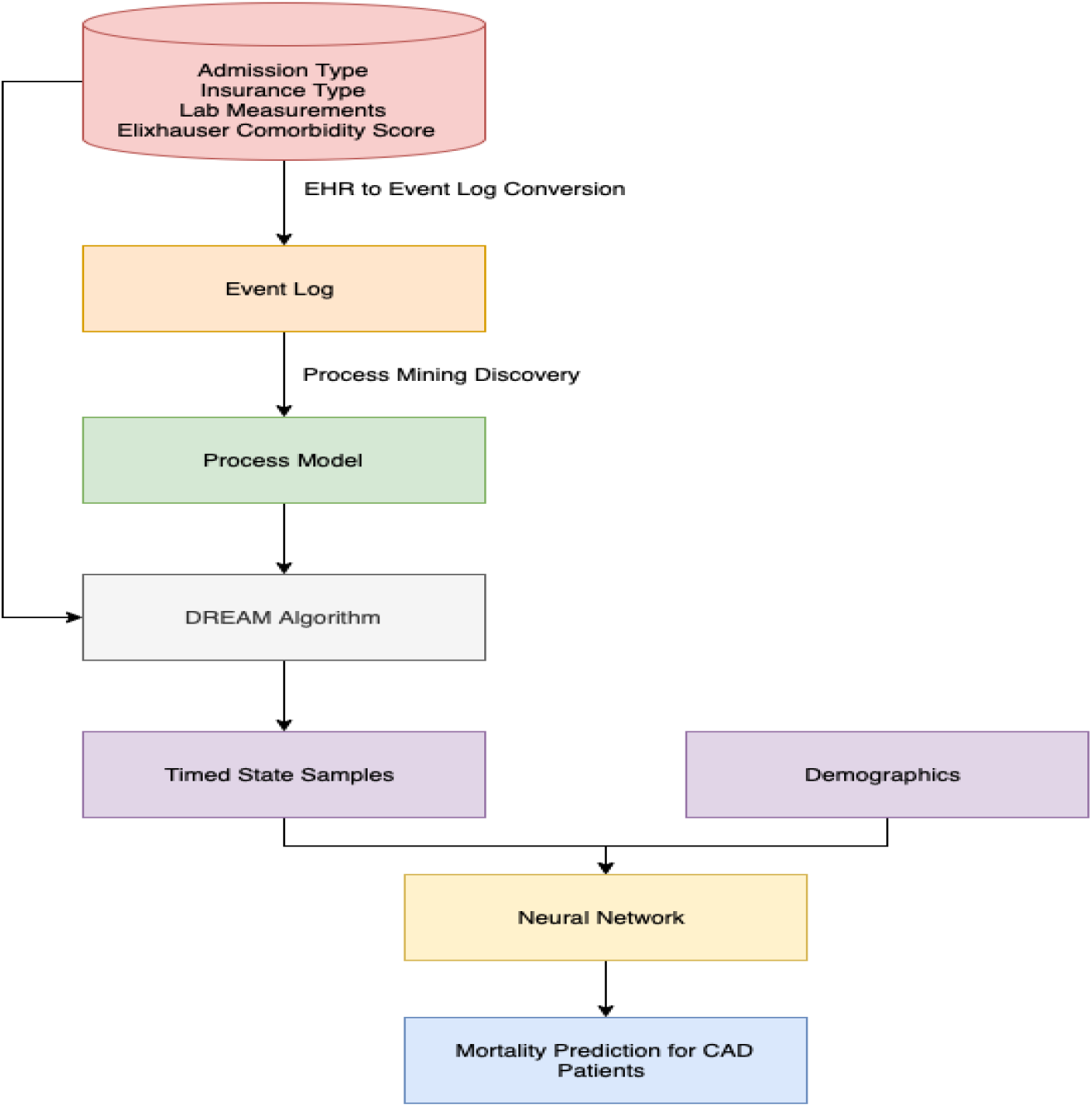
Framework for Mortality Prediction in CAD Patients

### 3.1. Feature Selection

The MIMIC III dataset was used for our trials. We only consider patients with at least one admission prior to the current one. Table 1 lists features extracted from MIMIC III [11]:

**Table 1:**
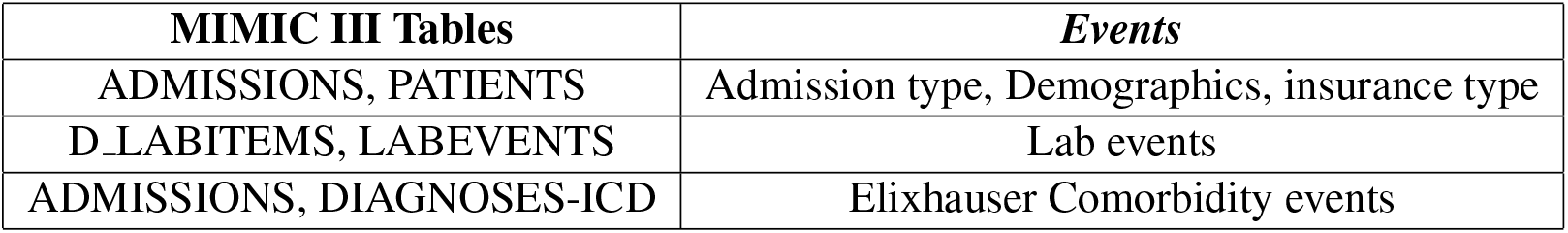
Converting MIMIC III tables to Event Logs.

1. *Admission Type:* Emergency, Elective, Urgent.
2. *Insurance Type:* Medicaid, Medicare, Private, Self Pay, Government.
3. *Lab Items:* Creatinine Kinase, Troponin T, HDL-Cholesterol, LDL-Cholesterol.
4. *Age, and gender*.
5. *Elixhauser Comorbidities*.

### 3.2. Converting EHRs to Event Logs

Event logs report the sequence of events with timestamps of when events occur. Events include patient *careflows* like admission, diagnosis, lab measurements, etc. Event logs contain 20 distinct events extracted from patient EHRs. 3 are admission types, 5 are insurance types, and 4 are lab measurements. The remaining distinct events belong to Elixhauser Commodities. Each event is unique and has a timestamp indicating when the event occurred. Timestamps for comorbidity events are set to the discharge time of the corresponding hospital admission. Events in a trace occur sequentially. To maintain their order, we delayed comorbidity events with the same timestamps by adding 1 ms to them.

### 3.3. Predictions

DREAM was used to predict whether a hospital discharges a CAD patient or the patient dies. DREAM replays the event logs of a process model and generates time-related information, referred to as “timed state samples”. Timed state samples and demographics like age and gender, are fed to the dense NN.

The NN architecture consists of the following: time state samples are input to a unique branch of one hidden layer with 200 neurons. After the first layer, a dropout rate of *DO* = 0.2 is applied for regularization. Demographics are then fed into a single hidden layer of 20 neurons followed by a dropout rate of 0.2. These layers are concatenated into a subsequent layer containing 90 neurons with a dropout rate of *DO* = 0.2. The architecture for NN is shown in Fig. 2

**Figure 2:**
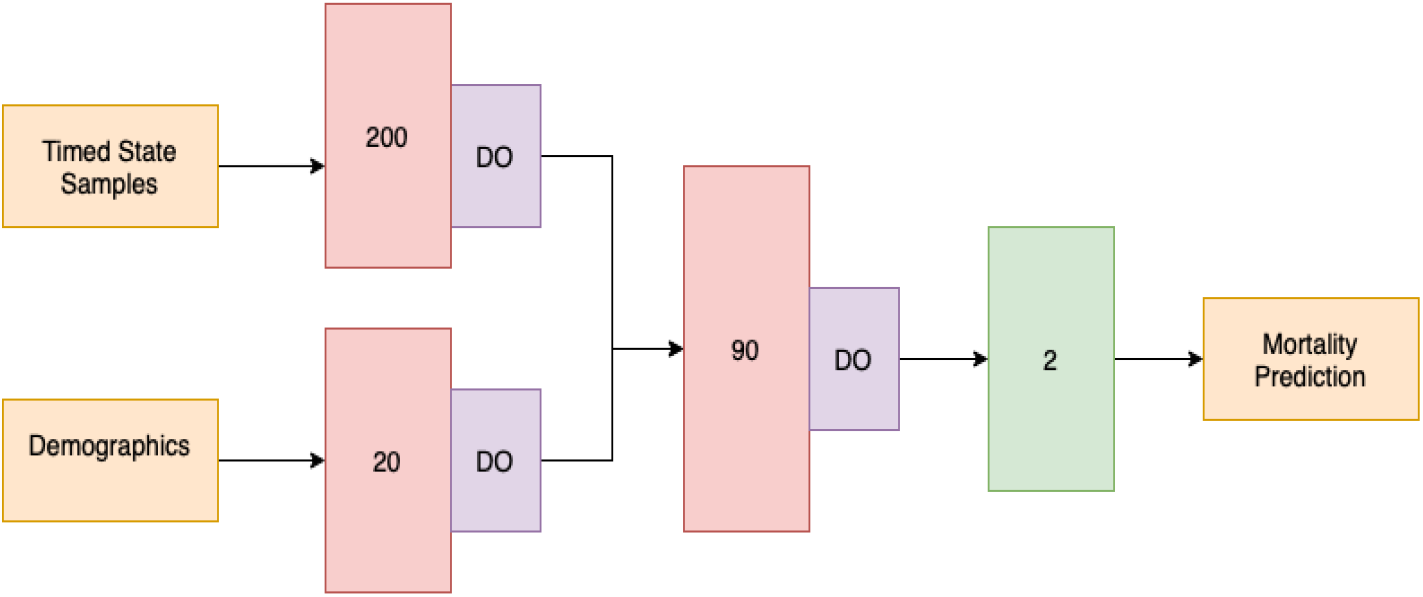
Neural Network Architecture

### 3.4. Baseline Models

The process mining/deep learning approach is compared to several baseline models to determine effectiveness. These models were constructed using machine learning algorithms and utilized the same variables. The performance of our proposed approach was compared to that of the machine learning algorithms. Machine learning algorithms included Support Vector Machine (SVM), Decision Trees, Random Forest (RF), and XG-Boost. These models were trained using a grid search to optimize model parameters. AUC of the validation cohort was used to select the best model.

## 4. Evaluation

Evaluation of the model to predict mortality for patients with CAD is discussed in this section. The dataset is described in the first subsection. The modeling setup is then discussed. Finally, results and comparisons to the baseline models are provided.

### 4.1. Dataset

MIMIC III (Medical Information Mart for Intensive Care) [11] is a large database of information on patients admitted to Beth Israel Deaconess Medical Center (BIDMC). Patient data includes vital signs, medications, laboratory measurements, observations and notes charted by care providers, fluid balance, procedure codes, diagnostic codes, imaging reports, hospital length of stay, survival data, etc. [11]. Data from 2,177 CAD patients were extracted from 46,476 total patients from ICU admissions in the MIMIC III database. The ICD-9 code was used for extraction.

Two data types were prepared. The first consisted of event logs of 20 unique events, including EHR information about patients like admission type, insurance type, lab items, and Elixhauser scores. The second consisted of patient demographic data like age and gender.

### 4.2. Setup

A dataset of 2,177 patients was randomly split into two sets, 69% for training (1503 patients) and 31% for testing (674 patients). To discover a process model and train the NN, the training set was also randomly split into train and validation sets using an 80*/*20% ratio respectively. The train and validation set were used to select the best model while the test set was used to evaluate model performance.

The NN was trained for 300 epochs using a batch size of 56 with a learning rate of 5e-4, and optimized using the RMSprop [12] algorithm. The AUC score was used as the metric to indicate the probability that a classifier ranks a randomly chosen positive instance higher than a randomly chosen negative one. The AUC score estimates performance better than other common classification performance metrics [13]. Higher AUC scores indicate the model better distinguishes between patients discharged and patients who die. A 95% confidence interval (CI) for the AUC score was also calculated using DeLong’s method [14].

### 4.3. Results

Results for proposed and baseline models are summarized in table 2. The AUC score for the proposed approach was higher than all baseline models on the test set. The CI of the proposed model was also the narrowest of all models.

**Table 2:** Summary of Results for Proposed and Baseline Models.

| Models | AUC, CI |
| --- | --- |
| Proposed model | 87.1, [83.1-91.3] |
| SVM | 82.3, [75.1-86.3] |
| Decision Tree | 80.5, [72.1-87.3] |
| RF | 84.2, [79.1-90.1] |
| XGBoost | 83.2, [78.4-89.3] |

Process mining is often preferred over state-of-the-art machine learning methods because it discovers a PN that graphically depicts (i.e. visualizes) all processes patients go through in a medical system. This visualization aids in interpreting results. Process mining also uses patients’ medical histories from prior hospital admissions. Lastly, process mining, such as our framework, can incorporate time-related information from events and timed state samples as features of an NN. However, the traditional methods are not capable of implementing them.

Our proposed approach is limited by the amount of information available in the patient’s medical records. Small clinics and hospitals might have limited or no patient histories. This method would also exclude new patients and those patients with no prior admissions to the current hospital visit. Moreover, hospitals tend not to share patient data across networks. Thus, techniques for handling low sample sizes could mitigate some of these limitations [15]. Also, the proposed approach works better in large hospital networks.

## 5. Conclusion

The proposed framework in this paper uses a process mining/deep learning-based approach to predict the mortality of patients diagnosed with CAD. The AUC score of 0.871 indicates improved performance compared to traditional machine learning models. The results also demonstrate that medical history and time-related information from events are important to predicting and evaluating the risk of death in CAD patients. Future work will explore other modeling techniques to increase performance in predicting mortality for CAD patients.

CAD patients are at high risk of death, therefore predicting patient outcomes more quickly would allow medical stakeholders to expedite treatments to save a patient’s life or to triage and reallocate resources to other patients. Our prediction framework can help medical teams make better decisions related to effective medical resource allocation that can increase the life expectancy of CAD patients.

Moreover, the framework’s ability to incorporate time-decayed event data into the neural network significantly enhances its predictive power, offering a more nuanced understanding of patient trajectories. This approach not only improves accuracy but also provides valuable insights into the progression of CAD, which can be critical for developing targeted interventions. By addressing limitations such as data completeness and cross-institutional variability, the framework can be further refined and adapted to a broader range of clinical scenarios, ultimately contributing to better patient outcomes and more efficient healthcare systems.

## Data Availability

All data produced are available online at:
https://physionet.org/content/mimiciii/1.4/

## Acknowledgment

The authors would like to thank all contributors of MIMIC-III for providing the public EHR dataset.

